# BUSINESS CONTINUITY PLANS FOR HOSPITAL PHARMACIES: A DELPHI CONSENSUS

**DOI:** 10.1101/2025.11.11.25339966

**Authors:** Ferdinand Le Bloc’h, Laurence Schumacher, Niels Kessler, Pierre-André von Zeerleder, Frank Calcavecchia, Pascal Bonnabry, Nicolas Widmer

## Abstract

**Objectives:** Business continuity plans (BCPs) are vital for hospital pharmacies (HPs) seeking to maintain essential operations during crises. Although generic BCPs exist, we found no guidelines for HP BCPs. We sought to use an expert consensus methodology to develop specific recommendations for BCP content for HPs.

**Methods:** An expert panel (comprising hospital chief pharmacists, deputies or quality managers) conducted a two-round Delphi study. Participants were recruited from civilian HPs in Switzerland and from two civilian and one military HP in each of Austria, Belgium, France, Germany and Italy. Recommendations were structured around the ‘all-hazards’ framework and practical examples and were developed based on the literature and investigators’ experiences in disaster simulation. A five-member, multidisciplinary steering committee refined the recommendations before each round of voting. Experts rated recommendations on a scale from 1–9, and the RAND/UCLA method was used to determine consensus. In Round 2, recommendations without consensus in Round 1, newly proposed recommendations, and recommendations with comments in Round 1 requiring rephrasing were submitted.

**Results:** Of 71 experts contacted, 24 participated, with 18 responding in Round 1 (75% response rate) and 15 responding in Round 2 (83% response rate). Most experts came from Switzerland (78%) and civilian hospitals (94%). Half were chief pharmacists and only 17% had no BCP available in their HP. In Round 1, 41 recommendations were submitted; 100% were validated. Eleven new recommendations were submitted in Round 2, including one new proposal and ten rephrased recommendations from Round 1. Experts had to select their preferred version. The new recommendation was validated, and all the rephrased recommendations were preferred. The final list comprised 42 recommendations for developing BCPs for HPs.

**Conclusion:** Our consensus-based BCP guidelines provide practical recommendations for civilian and military HPs and will help them develop BCPs for future crises and major business disruptions.

**KEY MESSAGES:** *What is already known about this topic:*

- Hospital pharmacies (HPs) lack tailored guidance for their business continuity plans (BCPs) for dealing with major disruptions.

*What this study adds:*

- This study presents 42 practical recommendations and examples of their use for developing BCPs for civilian and military HPs.
- The recommendations draw on a literature review, multidisciplinary perspectives and validation via a rigorous Delphi process involving an international expert panel of civilian and military pharmacists.

*How this study might affect research, practice or policy:*

- The recommendations aim to promote and support HPs’ planning processes, which should form a baseline for crisis preparedness and system resilience.
- The recommendations could eventually serve as a reference tool for all HPs and be adopted into quality policy certifications.

## INTRODUCTION

Disasters and other crises can seriously disrupt hospital operations. Direct physical damage to healthcare facilities and infrastructure can temporarily interrupt patient care^1^, while shortages of staff or essential medical products can delay or limit treatment^2^. Additionally, the increasing number of cyberattacks perpetrated on Europe’s hospitals represents a constant threat, with such potentially severe consequences as compromised patient safety, data breaches, psychological impacts and staff burnout^3^. Lastly, climate change further increases the likelihood of natural disasters^4^.

Nevertheless, healthcare providers, including hospital pharmacies (HPs), must always be able to maintain a minimum level of activity and ensure the provision of ‘routine’ care^5^. Ensuring the *business continuity* of essential services is a major step towards crisis preparedness and management^6,7^. For pharmacists, this involves ensuring the provision of services such as medicine distribution, safeguarding cold chain, or manufacturing sterile pharmaceutical preparations. Business continuity plans (BCPs) can assist in this, as they help define every measure that must be taken in the event of severe business disruption^8,9^. BCPs form part of broader risk resilience strategies^10-12^. They serve as guides for maintaining operational capabilities and restoring normal services in accordance with predefined objectives^10^. BCPs contain vital information on how to respond quickly to acute disruptions to normal business activities, offering solutions to reduce the likelihood or duration of those disruptions, limit the impact of specific incidents, protect priority activities and ensure the availability of adequate resources^6,10-12^. The planning process itself is an essential step in crisis preparedness and business continuity management (BCM)^7,11,12^. It must be accompanied by certain inherent human capacities for adaptation (managerial commitment, staff engagement, knowledge, innovation and training)^6^.

However, to the best of our knowledge, there were no formal recommendations regarding the necessary content for HPs’ BCPs. Several professional organisations^13-15^ have developed guides for creating and managing BCPs for community pharmacies. These guides could not be applied to hospital environments without adaptation, however. Any individual pharmacy or healthcare institution should tailor its BCP to its own unique context and local risks^6,8^. Nevertheless, being able to develop their planning within a common overarching framework would enable HPs to adopt the essential concepts of BCM and improve their preparedness for major disruptions. Although methodological guides^8,12^ and generic business continuity standards^10^ exist, none specifically addresses the particularities of HPs. The complexity and growing frequency of crises, their interdependence with other healthcare stakeholders and the absence of standards specific to HPs highlight the need for expert recommendations in this field^12^. Therefore, using an expert consensus methodology, this study’s primary aim was to establish a series of recommendations to support civilian and military HPs developing or improving their BCPs.

## METHODS

### Study design

We conducted a two-round Delphi study. The Delphi consensus method is an iterative research approach that solicits anonymous feedback from a group of experts on the subject matter using several rounds of questionnaires to achieve agreement on a particular issue^16^. Our study comprised three distinct groups, each with specific functions: investigators (conducting the study and formulating recommendations on BCPs for HPs), a steering committee (amending, correcting and approving those recommendations) and the Delphi expert panel (establishing consensus on the statements).

### Recommendation phrasing

Investigators phrased recommendations based on their findings in the existing literature (see references in Table S3) and their experiences in simulation exercises^17^. Each recommendation was supplemented by references and practical examples of HP activities. If a recommendation’s phrasing was significantly different from the literature, investigators signalled this with the suffix ‘EO’ (experts’ opinion). Recommendations were presented in four chapters under the ‘4Rs’ categories of risk Reduction, Readiness, Response and Recovery, a framework that follows the ‘all-hazards’ approach for preparing for major events and emergency situations^18^. Before each round of voting, recommendations were submitted to the steering committee, composed of five business continuity specialists from Switzerland: the chief pharmacist of a tertiary university hospital, the chief of the Swiss Armed Forces Pharmacy, the head of a hospital cybersecurity unit, the crisis and risk management advisor of a Swiss canton and the head of a hospital prevention, safety and security department. During a single meeting with the investigators, they discussed each recommendation and, if required, rephrased, removed or proposed additional recommendations based on their expertise. The final approved list of recommendations was then submitted to the expert panel for each round of voting.

### Expert panel

Expert panels must have a comprehensive knowledge of the topic at hand^19^, i.e. HPs operations; thus, our panel comprised hospital chief pharmacists or their deputy or departmental quality manager (whichever chief pharmacists preferred). Chief pharmacists from all of Switzerland’s hospitals were invited to participate, as were chief pharmacists from two civilian hospitals and one military hospital (from the Military Medical Corps Worldwide Almanac) in each of the neighbouring countries of Austria, Belgium, France, Germany and Italy. In total, 71 potential experts received an email invitation and a reminder to participate 1 month and 7 days before Round 1 voting, respectively. Experts who declined or did not respond were excluded. Given the number of potential experts, the high drop-out rates expected during Delphi processes and the desired balance between Swiss and foreign experts, a panel size of 30 experts was targeted^19,20^.

### Delphi rounds

Participating experts received an email inviting them to complete an online questionnaire containing the recommendations and to rate each one on a scale from 1–9 (1 = strongly disagree; 5 = neutral; 9 = strongly agree). They were able to add comments to qualify, correct, rephrase or remove recommendations, clarify an idea, request clarification or propose new recommendations. Questionnaires had to be completed within 4 weeks, with email reminders sent out 14 and 7 days before the end of Round 1. Eventually, a fifth week was accorded to maximise the response rate.

We used the RAND/UCLA appropriateness method to determine whether the panel had properly validated each recommendation^21^. This required the recommendation being accepted by the panel without dissent. Acceptance was measured using the median rating. Dissent was measured by comparing the distribution of ratings for a given recommendation with its symmetry-adjusted distribution. Briefly, each recommendation’s *Interpercentile Range* (IPRi) was compared to its *Interpercentile Range Adjusted for Symmetry* (IPRAS), which considers the asymmetry of rates by taking into account the distance between the IPRi and the scale midpoint (here, 5). The IPRAS is calculated as follows: IPRAS = IPRr + (AI ^*^ CFA), where IPRr is the perfect IPRi required for disagreement in case of perfect symmetry (found to be 2.35), CFA is the *Correction Factor for Asymmetry* (found to be 1.5) and AI is the *Asymmetric Index* (calculated as 5-IPCRP). IPCRCP is the *IPRi Central Point* and is calculated as (30% quantile + 70% quantile)/2. If a given recommendation’s IPR (IPRi) is larger than its IPRAS, there is dissent among the experts.

Recommendations submitted in Round 1 were considered validated by the panel if their median rating was ≥ 7, without dissent, and rejected if their median rating was < 7, without dissent. Recommendations with dissent were edited and resubmitted in Round 2, regardless of their median.

Recommendations with dissensus in Round 1, those validated in Round 1 but which the steering committee considered required relevant rephrasing after experts’ comments, and new recommendations proposed by the experts in Round 1 were all submitted for rating in Round 2. For each of these, experts received a personal report indicating their Round 1 rating and the panel’s median rating. The Round 2 questionnaire had to be completed within 3 weeks, with an email reminder sent out 7 days before the deadline. Round 2 used the same rating method, but when a recommendation validated in Round 1 was resubmitted with new phrasing, experts had to indicate their preferred version. The version with the highest median rating and (if the medians were equal) the narrowest IPRi was retained.

## Data analysis

Delphi rounds were conducted using the Research Electronic Data Capture platform (REDCap^®^, version 13.5.1). Raw data were exported into Microsoft Excel^®^ 2013 software (Microsoft Corporation, Redmond, WA, USA). Descriptive statistics (median, 30% and 70% quantiles, IPRCP, AI, IPRi and IPRAS) were calculated for each recommendation.

## RESULTS

We contacted 53 Swiss and 18 foreign experts (Figure 1), representing 75% and 25%, respectively, of the potential panel, with 66 (93%) civilian and 5 (7%) military experts. Of these 71 experts, 24 agreed to participate (34% acceptance rate), 4 declined and 45 did not respond. However, only 18 responded to the Round 1 questionnaire (75% response rate), with 15 responding to the Round 2 questionnaire (83% response rate). Table 1 presents the experts’ sociodemographic characteristics.

**Table 1:** Delphi experts’ sociodemographic characteristics.

| Characteristic – n (%) | Round 1 | Round 2 |
| --- | --- | --- |
| **Experts** | 18 (100) | 15 (100) |
| **Sex** |  |  |
| *Female* | 10 (56) | 7 (47) |
| *Male* | 8 (44) | 8 (53) |
| **Country** |  |  |
| *Switzerland* | 14 (78) | 11 (73) |
| *France* | 2 (11) | 2 (13) |
| *Belgium* | 2 (11) | 2 (13) |
| *Austria* | 0 (0) | 0 (0) |
| *Germany* | 0 (0) | 0 (0) |
| *Italy* | 0 (0) | 0 (0) |
| **Type of hospital** |  |  |
| *Civilian* | 17 (94) | 14 (93) |
| *Military* | 1 (6) | 1 (7) |
| **Position in the hospital pharmacy** |  |  |
| *Chief pharmacist* | 9 (50) | 7 (47) |
| *Deputy chief pharmacist / Management team member* | 6 (33) | 6 (40) |
| *Quality assurance manager* | 3 (17) | 2 (13) |
| **Years of experience in hospital pharmacy** |  |  |
| *< 5 years* | 0 (0) | 0 (0) |
| *5–10 years* | 4 (22) | 3 (20) |
| *10–15 years* | 2 (11) | 2 (13) |
| *15–20 years* | 5 (28) | 4 (27) |
| *> 20 years* | 7 (39) | 6 (40) |
| **Activities carried out in the hospital pharmacy (multiple answers possible)** |  |  |
| *Logistics* | 17 (94) | 14 (93) |
| *Clinical pharmacy* | 17 (94) | 15 (100) |
| *Drug manufacturing* | 14 (78) | 11 (73) |
| *Clinical trials* | 12 (67) | 10 (67) |
| *Quality control* | 10 (56) | 8 (53) |
| *Parenteral nutrition manufacturing* | 5 (28) | 4 (27) |
| *Sterilisation* | 1 (6) | 1 (7) |
| *Radiopharmacy* | 1 (6) | 1 (7) |
| **Training in business continuity management** |  |  |
| *Yes, I have undergone training with certification* | 0 (0) | 0 (0) |
| *Yes, I have undergone training but without certification \** | 2 (11) | 2 (13) |
| *No* | 16 (89) | 13 (87) |
| **Does the hospital pharmacy have a BCP?** |  |  |
| *Yes* | 6 (33) | 5 (33) |
| *Development is ongoing* | 9 (50) | 8 (53) |
| *No* | 3 (17) | 2 (13) |
| **In the last 5 years, has your hospital suffered a major event leading to an interruption of its activities (with the exception of COVID-19)? \*\*** |  |  |
| Yes | 2 (11) | 2 (13) |
| No | 16 (89) | 13 (87) |

\*Training reported by experts: Certificate of Advanced Studies in Medicines and Medical Devices in Emergencies and Disasters or personal investment.
\*\*Events reported by experts: Staff shortage (n = 2), Severe drug shortage (n = 1), Major stock-outs (n = 1), Financial crisis (lack of liquidity) or political crisis (lack of leadership) (n = 1), Cyber-attack (n = 1), Unavailable telecommunications (n = 1), Failure of major electronic equipment (n = 1).

**Figure 1.**
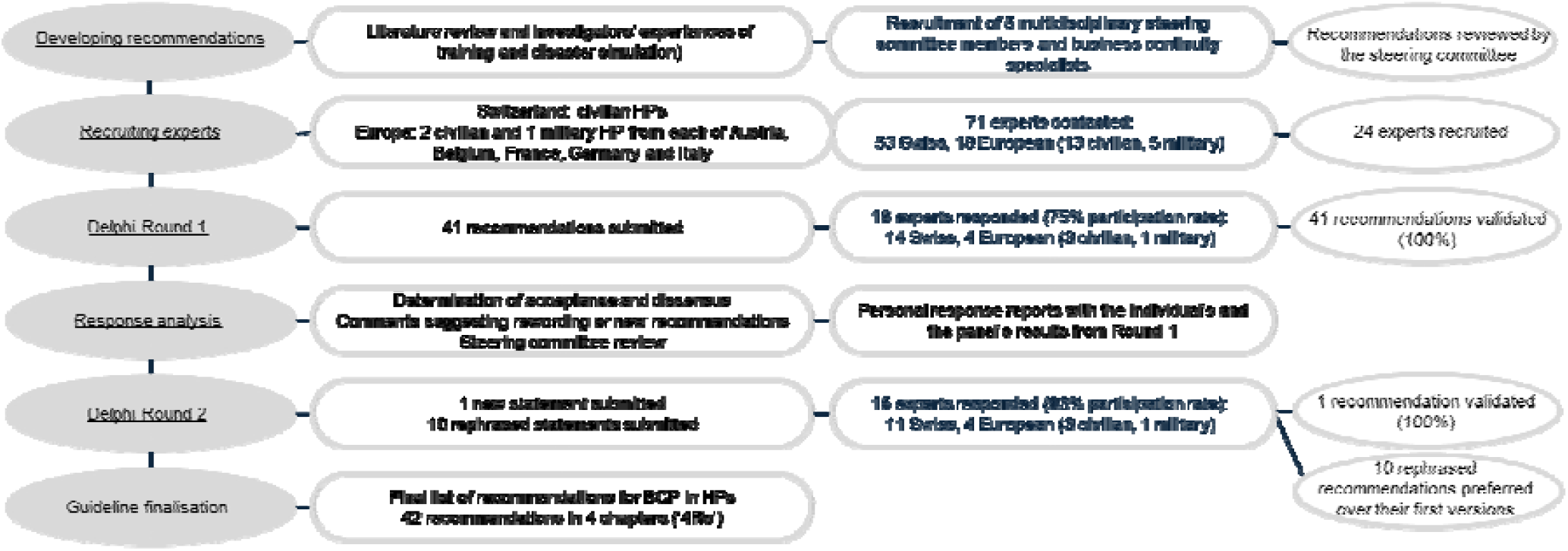
Delphi flowchart. ‘4Rs’ = risk Reduction, Readiness, Response and Recovery; BCP = business continuity plan; HPs = hospital pharmacies.

All the 41 recommendations were validated according to our method (median rating ≥ 7, no dissent). Only one new recommendation was suggested, regarding coordination with the HPs surrounding a given hospital. Rephrasing was proposed for a variety of recommendations and then analysed by the steering committee. Round 2 occurred two months later and lasted three weeks. Two experts provided their answers late, but within two weeks of the deadline, and were nevertheless considered. Eleven recommendations were submitted: one new and ten resulting from Round 1 rephrasing. The new recommendation was validated (median rating ≥ 7, no dissent), as were all those with proposed rephrasing, which were all preferred over their initial versions. Questionnaires and results from rounds 1 and 2 are presented in supplementary Tables S1 and S2, respectively.

The final list of 42 recommendations is available in Supplementary Table S3. Recommendations are presented using the ‘4Rs’ framework’s categories^18^ of risk Reduction, Readiness, Response and Recovery, with the chapters containing 26 (62%), 4 (9%), 7 (17%) and 5 (12%) recommendations, respectively.

## DISCUSSION

We developed a series of practical recommendations to provide HPs with simple, rigorous, consensus-based guidelines for creating BCPs. Recommendations were drawn from the existing literature and the multidisciplinary expertise of the study’s investigators and a steering committee, which included both pharmaceutical and non-pharmaceutical professionals. The well-established Delphi methodology, involving experts with extensive knowledge of HP management, was used to reach consensus, resulting in reliable and valid guidelines. This approach helped bridge the gap between theoretical best practice and real-world applicability in HP settings. The RAND method for reaching consensus was chosen based on previous studies^22^ and its validated statistical calculation methodology^21^. This helped create a robust framework that addressed the multiple challenges facing HPs in today’s healthcare landscape.

The study achieved a significant participation rate and strong engagement, with response rates of 75% (n = 18) in Round 1 and 83% (n = 15) in Round 2. The panel was smaller than targeted but similar to previous Delphi studies and included diverse expertise^19^. It was predominantly composed of experts from Switzerland (78%) and from civilian HPs (94%), reflecting our recruitment strategy. Four foreign experts (22%), including one military pharmacist, participated in both rounds. Most of the panellists were chief pharmacists (50%), had a long experience in HPs (67% had ≥ 15 years in the field) and led HPs performing multiple activities. This extensive expertise in HP management was essential to ensuring the relevance of our recommendations^23^. Notably, although none of our experts was formally certified in BCM, there were high rates of BCPs in place (33%) or in development (50%). This could be attributable to the current global context of uncertainty (e.g. cybersecurity threats, the Russo-Ukrainian war, supply issues, drug shortages)^3,24^. This contrasts with the 2018 survey by Schumacher *et al*.^25^, which found that only 27% of European HPs had disaster plans or standard operating procedures for crises. This difference in preparedness may also reflect lessons learned from the COVID-19 pandemic, as HPs with prior experiences of disasters are more likely to have disaster plans^25^.

All of Round 1’s 41 recommendations were validated according to the RAND method, as was the new recommendation in Round 2. None was rejected or faced dissensus. This may reflect the experts’ real needs for guidelines, as no specific BCP standard for HPs was available. It could also be linked to the concrete examples used to illustrate each recommendation, which can sometimes otherwise seem too generic and not very applicable at first glance. Finally, this could also be the result of the investigators and steering committee’s initial recommendation phrasing, which was a synthesis of the literature and their multidisciplinary experiences. Round 2 mainly involved rephrasing and fine-tuning recommendations to ensure they were comprehensive, practical and applicable for any future HP users.

Despite the high level of consensus achieved in Round 1, critical comments attached to several recommendations led to relevant rephrasing. The challenges of resource allocation were recurrent, including back-up materials, alternative premises and dedicated staff for implementing BCPs. Recommendation no. 2 (Table S1) emphasised the need to dedicate sufficient human, material, financial and time resources to resilience and preparedness. We did not provide examples with figures (e.g. full-time equivalents or number of back-up devices) as this depends on local factors such as an HP’s size or local predispositions to resilience. However, HP managers often lack the authority to secure these resources, requiring the hospital board’s agreement on funding and priorities (recommendation no. 1, Table S2). Experts commented on recommendation no. 27 (BCP training and education), highlighting the time and organisational constraints put on exercises. However, disaster training is indispensable to developing and validating plans, improving stakeholder familiarity and testing organisational resilience^5-9,11^. Simulation exercises are widely used for crisis preparedness^26^, and some require little equipment, time or money^27^. Nevertheless, in accordance with the experts’ comments, a reduction in simulation training frequency was proposed and unanimously preferred in Round 2 (Table S2). Finally, the panel’s new Round 1 recommendation to establish mutual aid agreements between HPs was widely accepted in Round 2 (median rating = 8). As above, Round 1 comments noted gaps in governance that limited HPs’ ability to formalise such agreements (recommendation no. 22). This underscored the need for hospital boards to empower BCM staff with resources and decision-making authority^12^.

BCPs must balance general disaster preparedness with the flexibility to adapt to specific events^6,11^. Preparing for events is crucial to reduce their likelihood and vulnerability to them, particularly for healthcare systems^5,9,26^. The World Health Organization advocates an ‘all-hazards’ approach, which emphasises adaptable, generic principles for major events preparedness rather than hazard-specific plans that would generate unnecessary duplications of work^12,28^. Reflecting this, our recommendations were structured around the ‘4Rs’ framework^18^. Combining the ‘all-hazards’ approach with the principles of BCM allowed us to focus on actions that enhance preparedness for both general and specific local threats, enabling responses to a wide range of business disruptions^5,9,12^. Indeed, 71% of our recommendations focused on risk Reduction and Readiness because BCM requires a thorough understanding and analysis of organisational processes to identify major vulnerabilities and establish protective and back-up measures. Conversely, only 17% of our recommendations focused on risk Response, since most of the measures needed during a business interruption should have been identified in the Reduction phase. The Response phase should then guide crisis communication and the application of these measures^12^. Thus, we have provided the general crisis management principles for HP settings based on the relevant literature and investigators’ experiences^12,29^. Ultimately, using the ‘4Rs’ framework aims to provide a comprehensive BCP development process through the structured categorisation of the progression of a business disruption^9^.

Our findings have many potential practical implications, as specific BCP recommendations for HPs have so far been lacking. Our tool aims to enhance HPs’ understanding of BCM and their preparedness in a time of ongoing crises^3,24^. Since most of our experts were from Switzerland (78%), these recommendations are primarily tailored for that country. However, they remain relevant internationally thanks to the participation of European experts (> 20% of the panel) during the Delphi process. Differences in practices between countries should be considered, as some results may reflect the unique characteristics of a single healthcare system (e.g. Switzerland’s HPs do not manage blood products). However, incorporating European experts undoubtedly helped broaden the guidelines’ perspectives and applicability. Our recommendations aimed to be generic enough to adapt to local contexts and could serve as a foundational framework for HPs’ BCPs, regardless of location. Finally, these guidelines could also prompt updates in HP quality standards certifications, putting new emphasis on BCM.

The present study had some limitations. Every Round 1 recommendation was validated, potentially indicating a lack of specific BCM training among experts, although their extensive HP experience mitigated this concern. Additionally, all the references used to develop our recommendations—including certifications^10,30^—were cited and made available to the experts, who could consult them freely. The inclusion of just one military pharmacy expert limited input from these crisis management professionals, yet her numerous contributions during Round 1 were very valuable in refining our recommendations, adding to the expertise of another military pharmacist on the steering committee. As mentioned, most of the experts were from Switzerland, with supplementary participation by French and Belgian experts. There were no representatives from Germany, Austria or Italy. The geographical and linguistic proximity of these experts to the investigators may have facilitated their participation and completion of the questionnaires. However, the recommendation development process—including a literature review, crisis simulation insights and steering committee input—and the Delphi method’s promotion of group consensus^16^ likely minimised the impact of a less broadly international panel. Finally, all the rephrased recommendations proposed in Round 2 were preferred over their initial versions, suggesting that the other recommendations might also have benefitted from further refinement, but that comments concerning them might have been subjectively ignored by the steering committee.

## CONCLUSION

The present Delphi study resulted in simple, consensus-based guidelines for developing business continuity plans (BCPs) for civilian and military hospital pharmacies (HPs). The 42 recommendations developed by an international expert panel are accompanied by tangible examples and are intended to be practical and adaptable to local contexts. The guidelines follow a ‘4Rs’ structure (risk Reduction, Readiness, Response and Recovery) based on the World Health Organization’s ‘all-hazards’ approach. Future research should measure the impact of the guidelines’ deployment in HPs and their applicability across different European countries. The present work contributes to raising awareness about pharmacy business disruptions in hospital settings and will help improve preparedness and resilience for future crises.

## Data Availability

All data produced in the present study are available upon reasonable request to the authors.

## ACKNOWLEDGEMENTS AND AFFILIATIONS

The authors warmly thank all the experts for their valuable contributions and dedicated involvement in this Delphi study. The authors gratefully acknowledge the financial support of the Swiss Armed Forces’ Competence Centre for Military and Disaster Medicine, Bern, Switzerland. They declare that they have no competing interests other than those resulting from the study’s funding and no affiliations with the pharmaceutical industry.

## CREDIT AUTHORSHIP CONTRIBUTION STATEMENT

**Ferdinand Le Bloc’h:** Conceptualization, Methodology, Formal analysis, Resources, Writing - Original draft. **Laurence Schumacher:** Conceptualization, Methodology, Supervision, Writing - Review & Editing. **Niels Kessler:** Writing - Review & Editing. **Pierre-André von Zeerleder:** Writing - Review & Editing. **Franck Calcavecchia:** Writing - Review & Editing. **Pascal Bonnabry:** Conceptualization, Funding acquisition, Supervision, Writing - Review & Editing. **Nicolas Widmer:** Conceptualization, Funding acquisition, Supervision, Writing - Review & Editing

## ETHICS APPROVAL

The research protocol was presented to the Canton of Geneva’s Research Ethics Committee, which waived the requirement for approval.

## SUPPLEMENTAL MATERIAL

- Table S1 – Delphi Round 1 survey
- Table S2 – Delphi Round 2 survey
- Table S3 – Final list of recommendations for hospital pharmacy business continuity plans

## BCPs FOR HOSPITAL PHARMACIES: A DELPHI CONSENSUS

