## Supplemental Tables S1 and S2 for "BUSINESS CONTINUITY PLANS FOR HOSPITAL PHARMACIES: A DELPHI CONSENSUS"

SUPPLEMENTAL FILE

**Table S1** – Delphi Round 1 recommendation results for business continuity plans for hospital pharmacies

| **Statement number** | **Recommendation** | **Examples** | **Median (n = 18)** | **Dissensus** | **Rephrasing required** |
| --- | --- | --- | --- | --- | --- |
| **I. REDUCTION** | | | | | |
| **Business continuity policy** | | | | | |
| 1 | The BCP must clearly define the hospital pharmacy’s **business continuity objectives.** These objectives must be achieved through **activities and processes that are deemed critical** and must be defined in **coordination** with the pharmacy’s external partners (suppliers and recipients of services). | *Objectives specific to each entity. Guaranteeing that care units are supplied with the medicines on the hospital’s medicines list; maintaining the traceability of flows or stocks of medicines; maintaining a minimum in-hospital drug manufacturing capacity for certain products; guaranteeing pharmaceutical information support for hospital care units; maintaining training capacity for staff in continuing postgraduate education; guaranteeing employee protection and safety; ...* | 9 | No | Yes |
| 2 | Pharmacy department management must dedicate **sufficient resources** to developing, updating and improving the BCP. | *Assigning dedicated and/or trained staff; preparing job specifications; dedicating sufficient time; dedicating financial resources to replacing equipment or training staff about plans; ...* | 8,5 | No | No |
| 3 | Given the specific roles of the various professionals working in hospital pharmacies, it is recommended that the **person responsible for piloting the BCP have an overview** of every business process as well as **managerial decision-making powers**. A working group made up of representatives from each pharmacy sector/unit can also be proposed. | *Member of the management team, the hospital pharmacy quality assurance manager or a dedicated risk manager.* | 9 | No | Yes |
| 4 | Given the specific roles of the various professionals working in hospital pharmacies, it is recommended that **each BCP item be adapted to each pharmacy unit/sector**, within a common framework. | *The pharmacy’s diversity of tasks and activities (pharmaceutical assistance and clinical pharmacy, drug supply, drug distribution, manufacture of therapeutic products, management of clinical trials, etc.); diversity of computer software used (electronic patient records, stock management software, chemotherapy or parenteral nutrition manufacturing software, etc.); variable possibility of replacement depending on profession and staff qualifications (specific clinical activities, training for pharmacy assistants, regulatory requirements in terms of cleanliness and asepsis, etc.); materials and equipment vary depending on profession and staff qualifications (specific clinical activities, training pharmacy assistants, regulatory requirements in terms of cleanliness and asepsis, etc.); heterogeneous materials and equipment use, with variable requirements (automated drug cabinets, clean rooms and isolators, medical devices, etc.); different forms of collaboration and interdependence with other departments; …* | 9 | No | No |
| **Business impact analysis (BIA)** | | | | | |
| 5 | A **detailed analysis of the pharmacy’s activities** must be a **prerequisite** for developing a BCP. It must identify and **address all the hospital pharmacy’s activities** with the goal of establishing an exhaustive assessment of the criticality of those activities. | *Administrative activities (planning, invoicing, etc.); training (postgraduate training, teaching provided to medical and nursing teams, etc.); quality control activities; subcontracting external drug manufacturing; updating documentation; managing clinical trials; etc.* | 9 | No | No |
| 6 | Each activity’s requirements in terms of **human resources**, **equipment**, **information and communication systems**, **infrastructure** and **external dependencies** must be identified. | *Human resources: duties, qualifications required, minimum numbers of staff, etc. Equipment: storage and distribution robots, automated weekly pill dispensers, automated drug cabinets, scales and analysis equipment, laminar flow cabinets, shelving, refrigerators, insulated crates, etc. Information and communication systems: Telephones, computers, internet, software, online databases, etc. Infrastructure: Offices, buildings, changing rooms, airlocks, preparation rooms and required qualifications, ventilation, cold rooms, goods-in areas, storage areas and required dimensions, etc. External dependencies: the host hospital institution, care units, suppliers, equipment manufacturers, delivery and transport companies, ad hoc committees, laboratories, other hospital pharmacies, customers, etc.* | 9 | No | No |
| 7 | The BCP must **clearly identify critical hospital pharmacy activities and processes** and consider the links between them. These critical activities and processes must ensure that the pharmacy achieves its business continuity objectives. | *Medicine stock management; supply of medicines to specific care units; manufacture of specific medicines; management of medical waste; access to a pharmaceutical helpline; ...* | 9 | No | No |
| 8 | For each critical activity and process, the BCP must describe the **minimum level of service required**, the **minimum level of resources needed to achieve them** and the **maximum duration for which this minimum service** is acceptable. | *Minimum service required: reduction in the frequency of drug distribution; manufacture of chemotherapies exclusively; etc. Minimum resources required: number of pharmacy assistants, pharmacy technicians or pharmacists required; use of specific software; etc.* | 9 | No | No |
| 9 | The BCP must describe the **maximum duration for which each critical activity or process** **interruption** is acceptable. | *Maximum duration of interruption: service provision interrupted for 24 hours; billing interrupted for one week; clinical pharmacists’ visits interrupted for one month; pharmaceutical helpline interrupted for 30 minutes; etc.* | 8,5 | No | Yes |
| 10 | **A business impact analysis (BIA)** must be a **prerequisite** for developing a BCP and must identify the risks and threats to the continuity of the hospital pharmacy’s activities. These risks can be internal or external to the pharmacy. | *Possible risks and threats: disruption to drug supply; failure of certain critical installations; electrical or IT failure; telecommunications problems; human resources shortages; massive influx of patients; unavailability of premises; natural disasters; man-made disasters; etc. Risk analysis: using the FMECA method (Failure Modes, Effects and Criticality Analysis).* | 9 | No | Yes |
| **Risk prevention and mitigation** | | | | | |
| 11 | The BCP must include a **strategy for replacing staff** in key roles and involved in critical activities and processes, particularly in the event of a long-term crisis. This can be done using a **substitution list** or by **training staff** from different sectors/units internally to ensure interoperability. | *Replacing the pharmacy assistant responsible for critical care units; replacing pharmaceutical information hotline; ... Defining which staff need to know how to use an automated drug cabinet; defining who needs to be able to validate hospital drug manufacturing, etc.* | 9 | No | No |
| 12 | The BCP must include a strategy for **mobilising and reassigning staff** if necessary. This strategy must be developed in collaboration with the **institution’s human resources managers.** The strategy can **evolve over time** and differ depending on the situation and the duration of the crisis. | *Recalling staff on leave; changing working hours; calling on external resources such as staff from other hospital pharmacies, the army or civil defence forces; recruiting quickly through temporary employment agencies; mobilising pharmacy students; …* | 8,5 | No | Yes |
| 13 | The BCP must consider **staff safety** in the event of a crisis and give specific instructions to ensure it. | *Epidemiological protection measures (PPE, working from home, exposure of pregnant or breast-feeding women, etc.); physical protection (against fire, risk of a building collapse, physical aggression, physical exhaustion following overwork, etc.); psychological protection (from traumatic situations, mental exhaustion, etc.).* | 9 | No | No |
| **Infrastructure** | | | | | |
| 14 | The BCP must set out the possibilities for **replacing critical materials or equipment**, if they exist, or the potential alternatives in the event of their unavailability. | *Back-up computers; back-up refrigerators or those of other hospital departments; spare parts for robots; pallet transporters; etc.* | 9 | No | No |
| 15 | The BCP must identify the **critical IT software** used by the hospital pharmacy and describe the **solutions to be used** in the event of their failure. This work should be based on mapping the software and applications used by the hospital pharmacy. | *Critical software: Electronic patient records; stock management software; invoicing software; chemotherapy or parenteral nutrition manufacturing software; ...* | 9 | No | No |
| 16 | The BCP must identify the hospital pharmacy’s **critical IT data**, define how it will be **backed up** and describe **data recovery solutions**. | *Critical data: stock levels; list of suppliers; list of supplies; order forms; manufacturing and analysis protocols; pharmaceutical support documents; list of databases and associated passwords; plans and procedures; etc. Also: staff data; invoicing data; pharmacy bank account numbers; etc. Back-up methods: regular paper printouts; manual back-ups onto a hard disk; automated back-ups onto a computer or server; etc.* | 9 | No | Yes |
| 17 | The BCP must define available **alternative means of telecommunication**. These means must be known to all staff and/or be available during normal periods to facilitate their use in the event of the BCP being activated. | *Replacement telephones; internal paper-based messaging; transport or delivery staff that can deliver paper mail; physical movement between hospital departments or hospital buildings; using fax machines; etc.* | 9 | No | No |
| 18 | If the possibility exists, the BCP must describe **alternative premises that could house all or part of the hospital pharmacy**. It is recommended that these premises should be able to house, at the very least, the pharmacy’s stock of medicines, but also preferably staff workstations and the hospital’s manufacturing laboratories. | *Available space if storage robots need to be emptied; large off-site rooms if the pharmacy is unusable; warehouses; transportable clean rooms; hospital operating theatres; external laboratories such as another hospital pharmacy or a private company; etc. If necessary, do not forget medicine storage equipment (shelves, pallets, etc.) and electrical power for fridges/cold rooms.* | 8 | No | No |
| 19 | The BCP must describe the **local electrical network** and identify, if it exists, the **back-up network and its** **technical details**. It must also describe the **critical electronic devices and equipment** that must have priority access to this back-up network. | *Identifying emergency power outlets and continuous or intermittent power supply to them; equipment connected to emergency power; duration of the building generators’ autonomy; energy consumption of the pharmacy’s electrical appliances; the possibility of using external batteries for certain appliances; etc.* | 9 | No | No |
| 20 | The BCP must consider **other energy sources and options**, as well as the environmental working conditions in which the hospital pharmacy operates. | *Water used to manufacture hospital products; heating and maintaining storage room temperatures; necessary ventilation; ventilation of premises; ...* | 8,5 | No | No |
| 21 | The BCP must identify and describe the **procedures for accessing pharmacy premises** in the event of a security badge malfunction. The same applies to accessing other **critical areas requiring security badges** (narcotic medicine cabinet, etc.). | *Identifying how the host hospital’s security badge system works; identifying critical access doors; identifying door opening procedures using a physical key; contact persons in the event of security badge malfunction; consideration of stock alarm triggering procedures; possibility of calling in security guards; etc.* | 9 | No | No |
| **Collaboration with partners** | | | | | |
| 22 | The BCP must identify the hospital pharmacy’s **main external partners** from its BIA. It must define with them the possible **adaptations to operations** that can be made in the event of a partial or total breakdown in operational capacity. It is recommended that these adaptations **be defined in collaboration** with these partners. | *Collaborating with other hospital pharmacies on taking over some activities, such as hospital drug manufacturing, lending equipment, supporting human resources, taking over some drug storage or orders, etc. Rescheduling clinical activities in some care units; borrowing hospital equipment or premises; adapting ordering procedures with drug suppliers; targeted support for IT teams to restore critical data; etc.* | 8,5 | No | No |
| 23 | The **analysis of IT risks** must be a **prerequisite** to the development of the BCP and must be carried **out in collaboration with the host hospital institution’s IT teams** or with the help of its IT subcontractors. IT back-up procedures described in the BCP **should be** **defined by these IT teams** so that they coordinate with their own plans. | *IT failure analysis; cyber-attack risks; software and data mapping; defining data back-up and recovery procedures; prioritising support to critical IT applications; borrowing IT equipment; ...* | 9 | No | No |
| **Specific scenarios** | | | | | |
| 24 | It is recommended that **detailed procedures** be established to deal with the **most critical risks** to the hospital pharmacy. These can be risks with a high probability of occurring or risks that would have major impacts on critical activities. These procedures **must be listed in the BCP**. | *Massive inflows of patients; major staff shortages; major malfunctions of hospital drug manufacturing equipment (clean rooms); power cuts; major IT failures (breakdowns, cyber-attacks, etc.); loss of premises (flood, fire); etc.* | 9 | No | No |
| 25 | The BCP must define a **list of essential medicines and products**, setting their levels of criticality and minimum stock levels. Collaboration with the care units concerned is recommended to determine this list and the required quantities. | *Classifying care units by levels of criticality, physiological system, ATC code, etc. Based on the needs of the care units concerned, their clinical importance, the difficulties replenishing them, the need for antidotes, the CBRN risks, etc.* | 9 | No | No |
| **II. READINESS** | | | | | |
| **Theoretical and practical training** | | | | | |
| 26 | Hospital pharmacy staff **must be trained about BCPs**. It is recommended that the training methods used, the levels of knowledge required and the minimum frequency of that training be described in advance. | *Examples: training in technical procedures such as chemotherapy manufacturing without computers or ventilation; drug ordering without appropriate software; pharmaceutical support for departments without access to certain databases; etc. Methods: types of training (theoretical or practical, partial or complete, etc.); target audiences (pharmacists or pharmacy assistants, team leaders, users of certain IT equipment or software, etc.); frequency of training; staff responsible for training (BCP manager, hospital or external service provider, etc.); levels of knowledge required and/or sought (first response in the event of a crisis, such as the BCP’s location or the first point of contact; a perfect understanding of how degraded equipment operates, such as automated drug cabinets, or how procedures such as telephone pharmaceutical assistance are maintained, etc.).* | 9 | No | No |
| 27 | **Critical BCP functions must be trained or tested at least once a year** or when the pharmacy undergoes a major reorganisation. Decisions on training methods are left up to the BCP manager; however, it is recommended to carry out **a simulation exercise** at least once a year and to practice performing critical activities **in a degraded mode of functioning or manually**. | *Critical topics: power cuts; testing IT back-up procedures; cyber-attacks; massive inflows of patients; staff substitution for critical tasks; staff understanding of crisis measures; ... Simulation exercises: quizzes; board games; tabletop exercises; emergency drills; functional exercises; full-scale exercises; virtual reality; exercises in collaboration with hospital care units; etc.* | 8,5 | No | Yes |
| **Continuous improvement** | | | | | |
| 28 | The BCP must be improved continuously. To do this, the BCP must have **defined assessment and updating procedures**; it must be reviewed and revised **after each exercise or real crisis event**. Document management software is recommended. | *Assessment procedures: Assessment types (tests, reviews, audits, post-incident reports); prerequisites; frequency of assessments; staff responsible for assessments; compliance with regulatory and/or legal requirements; follow-up of previous assessments; etc. Updating procedures: Frequency of updates (at regular intervals, after each training session or assessment, during major or significant changes within the hospital pharmacy, when the BCP is activated, etc.); staff responsible for updates; monitoring previous changes; communication procedures (communication channels, recipients, etc.).* | 8,5 | No | No |
| **Risk prevention and detection** | | | | | |
| 29 | The BCP must define the **means of monitoring and detecting risks** to hospital pharmacy activities. The individuals responsible for monitoring and detecting risks must be identified. | *A permanent monitoring unit; regular, planned discussions with other hospital pharmacies; tools for detecting risks of supply shortages; up-to-date information on regional risks; seasonal epidemic monitoring; collaboration with hospital and regional-level staff responsible for monitoring epidemics; ...* | 8 | No | No |
| **III. RESPONSE** | | | | | |
| **Crisis management** | | | | | |
| 30 | The BCP must define the **threshold indicators and procedures for activating the plan**. If necessary, decision-making stages can be specified depending on predefined situations or mechanisms. | *Triggering modes; staff authorised to activate plans; communication modes; priority contacts; indicators or threshold values defining levels for renewed decision-making (such as the number of staff missing, the unavailability of critical IT software or the duration of an activity interruption); etc.* | 8 | No | No |
| 31 | The BCP must describe the **composition of the hospital pharmacy’s crisis management team**. It is recommended that this team be composed of **hospital pharmacy management team members reinforced by staff with additional skills**. | *Minimum staffing numbers; required functions; leaders and their replacements in the event of an absence; role assignment, with or without job descriptions; actions expected from each stakeholder; defining governance of the various stakeholders; defining meeting places; …* | 9 | No | Yes |
| 32 | The BCP must refer to the use of appropriate **crisis management tools**. It is recommended that these tools be available in both electronic and paper formats. | *Management tools: event diary; situation chart; visual analysis of the situation; risk anticipation chart; staff diary and human resources management chart; problem tracking chart; chart for tracking new tasks or activities related to the crisis event; ... Tool accessibility: tool storage locations are known; tool use has been tested by the staff concerned; ...* | 8 | No | No |
| **Continuity management** | | | | | |
| 33 | The BCP must refer to **decision-support tools** for maintaining, adapting or suspending activities. These tools can also be used to monitor critical activities and processes. | *Monitoring essential pharmaceutical products; tracking human resources; monitoring drug orders and distribution; monitoring drug shortages; prioritising the computer software to be restored; ordering the interruption of activities according to the number of staff absent; ...* | 8 | No | No |
| 34 | It is **recommended that BCP solutions** for handling interruptions to pharmacy activities **be formulated** so that all pharmacy staff can easily understand and use them. | *Precise instructions for implementing strategies; clear identification and distribution of roles; identification of resources required for implementation; references to associated degraded procedures, if they exist; ...* | 9 | No | No |
| 35 | The BCP must define **requirements and procedures for the traceability and continuity of information**, particularly in the event of degraded modes of activity for processes with highly dynamic data. | *Traceability of incoming or outgoing stocks using a degraded Excel™ spreadsheet or manual system; the continuity of information regarding patients’ chemotherapies (protocols, information transmission to oncology teams, ...); monitoring the staff’s working hours (whether in normal or exceptional situations); decisions made with partners such as suppliers; ....* | 9 | No | No |
| **Communication** | | | | | |
| 36 | The BCP must identify and describe **communication procedures to be used in the event of a crisis**. This concerns communication both inside and outside the pharmacy. | *Staff responsible; preferred communication channel (telephone, smartphone messaging, website, in-person meetings, etc.); preprepared messages; up-to-date lists of useful contacts (hospital care units, other hospital pharmacies, hospital technical or IT departments, suppliers, etc.); up-to-date lists of staff; frequency of communication (depending on audience: every hour, every day, etc.); media/general public communication policy; etc.* | 9 | No | No |
| **IV. RECOVERY** | | | | | |
| **Coming out of a degraded mode of functioning** | | | | | |
| 37 | The BCP must define **procedures for coming out of the crisis situation and ending the plan**. | *Staff authorised to declare the end of the crisis; means of communication; priority contacts; ...* | 8 | No | No |
| 38 | For each critical activity and process, the BCP must set a **target timeframe for a return to normal activities**. This can be done by means of a **recovery plan** that defines the steps to be taken to recover from a degraded situation, identifying the activities and processes to be resumed according to the situation and available resources. | *A return to normal chemotherapy manufacturing volumes within three days; a return to the normal weekly drug replenishment frequency for every department; etc. Defined recovery times, levels of recovery, and the resources that need to be mobilised for that recovery.* | 8 | No | Yes |
| 39 | It is recommended that **staff be debriefed** after any exercise or activation of the BCP. It is recommended that the BCP define the **debriefing procedures**. | *Debriefings during and immediately after crisis situation; staff impressions and feelings; ... Procedures: Staff responsible for debriefings; referral to professionals if necessary (such as crisis managers or psychologists); ...* | 8 | No | No |
| **Return to normal** | | | | | |
| 40 | The BCP must include a reminder of the **aspects relating to a return to normal** after an exercise or activation of the plan. | *Procedural review if the new ‘normal’ situation has been modified as a result of the crisis situation (changes in suppliers, changes in drug supply flows to departments, new equipment or computer software, reorganisation of staff and their tasks, etc.); verification of the integrity and validation of critical equipment (automated drug cabinets, storage and distribution robots, ventilation, isolators, analytical tools, computer software, etc.); verification of staff qualifications; reassessment of certain processes in accordance with regulatory requirements (aseptic processes, etc.); …* | 8 | No | No |
| **Feedback on experiences** | | | | | |
| 41 | The BCP must include **tools for analysing crisis situations** to provide feedback and recommendations for improvement. | *Examples of items to analyse: Chronology of events; foreseeable or reducible disruptions; actions already in place to limit their impact; activities affected by the disruption; human, material or financial consequences; actions implemented during the disruption; difficulties encountered; shortcomings identified in the BCP’s ability to handle disruptions; analysis of actions that worked well and those that need improvement; financial impact of the emergency event and its management; planned improvements to the BCP; planned reporting.* | 8,5 | No | Yes |

N = 18 experts. BCP, business continuity plans ; BIA, business impact analysis ; IT, information technology; PPE, personal protective equipment.

**Table S2** – Results from Delphi Round 2 on the new suggested recommendation and on rephrased recommendations according to expert comments during Round 1.

| **Statement number** | **Rephrased recommendation** | **Rephrased examples** | **Experts’ preference – n (%)** | | | |
| --- | --- | --- | --- | --- | --- | --- |
|  |  |  | **1^st^ round version** | | **Rephrased statement** | **No preference or opinion** |
| **I. REDUCTION** | | | | | | |
| **Business continuity policy** | | | | | | |
| 1 | The BCP must clearly define the hospital pharmacy’s **business continuity objectives**. These objectives must be achieved through **activities and processes that are deemed critical** and must be defined in coordination **with the pharmacy’s external partners** (suppliers and recipients of services), including the direction of the hospital. | *Objectives specific to each entity.*  *Guaranteeing that care units are supplied with the medicines on the hospital’s medicines list; maintaining the traceability of flows or stocks of medicines; maintaining a minimum in-hospital drug manufacturing capacity for certain products; guaranteeing pharmaceutical information support for hospital care units; maintaining training capacity for staff in continuing postgraduate education; guaranteeing employee protection and safety; ..."* | 3 (20) | | 11 (73) | 1 (7) |
| 3 | Given the specific roles of the various professionals working in hospital pharmacies, it is recommended that the **person responsible for piloting the BCP have an overview** of every business process as well as **managerial decision-making powers**. A working group made up of representatives from each pharmacy sector/unit and critical equipment managers must also be set up. | *Member of the management team; the hospital pharmacy quality assurance manager; a dedicated risk manager; pharmacy technician responsible for the drug storage robot ; etc.* | 4 (27) | | 11 (73) | 0 (0) |
| **Business impact analysis (BIA)** | | | | | | |
| 9 | The BCP must describe the **maximum duration for which each critical activity or process** **interruption** is acceptable. It must also describe the actions to be taken depending on the incident **duration**. | *Maximum duration of interruption: service provision interrupted for 24 hours; billing interrupted for one week; clinical pharmacists’ visits interrupted for one month; pharmaceutical helpline interrupted for 30 minutes; etc.* | 3 (20) | | 11 (73) | 1 (7) |
| 10 | **A business impact analysis (BIA)** should be the **basis** **of the planning process** and must contribute to the **continuous improvement of the BCP**. This BIA must identify the risks and threats to the continuity of the hospital pharmacy’s activities; these risks can be internal or external to the pharmacy. | *Possible risks and threats: disruption to drug supply; failure of certain critical installations; electrical or IT failure; telecommunications problems; human resources shortages; massive influx of patients; unavailability of premises; natural disasters; man-made disasters; etc. Risk analysis: using analysis tools, e.g. the FMECA method (Failure Modes, Effects and Criticality Analysis) or the SWOT method (Strengths, Weaknesses, Opportunities, Threats).* | 3 (20) | | 12 (80) | 0 (0) |
| **Risk prevention and mitigation** | | | | | | |
| 12 | The BCP must include a strategy for **mobilising and reassigning staff** if necessary. This strategy must be developed in collaboration with the **institution’s human resources managers** as well as **the direction of hospital**. The strategy can **evolve over time** and differ depending on the situation and the duration of the crisis | *Recalling staff on leave; changing working hours; calling on external resources such as staff from other hospital pharmacies, the army or civil defence forces; recruiting quickly through temporary employment agencies; mobilising pharmacy students; …* | 2 (13) | | 10 (67) | 3 (30) |
| **Infrastructure** | | | | | | |
| 16 | The BCP must identify the hospital pharmacy’s critical IT data, define how it will be backed up and describe data recovery solutions. | *Critical data: stock levels; list of suppliers; list of supplies; order forms; manufacturing and analysis protocols; pharmaceutical support documents; list of databases and associated passwords; plans and procedures; etc. Also: staff data; invoicing data; pharmacy bank account numbers; sales or purchases contracts ; etc. Also : patient-related data, clinical trials data, etc. Back-up methods: regular paper printouts; manual back-ups onto a hard disk; automated back-ups onto a computer or server; etc.* | 2 (13) | | 12 (80) | 1 (7) |
| **II. READINESS** | | | | | | |
| **Theoretical and practical training** | | | | | | |
| 27 | **Critical BCP functions must be regularly trained or tested, ideally once a year** or when the pharmacy undergoes a major reorganisation. Decisions on training methods are left up to the BCP manager; however, it is recommended to carry out **a simulation exercise** and to practice performing critical activities **in a degraded mode of functioning or manually**, and to adapt the type of training **according to the public targeted (management team, operational staff).** | *Critical topics: power cuts; testing IT back-up procedures; cyber-attacks; massive inflows of patients; staff substitution for critical tasks; staff understanding of crisis measures; ... Simulation exercises: quizzes; board games; tabletop exercises; emergency drills; functional exercises; full-scale exercises; virtual reality; exercises in collaboration with hospital care units; etc.*  *Moment of training: induction program for new employees; part of an audit; major reorganisation of the pharmacy; planed schedule; …* | 0 (0) | | 15 (100) | 0 (0) |
| **III. RESPONSE** | | | | | | |
| **Crisis management** | | | | | | |
| 31 | The BCP must describe the **composition of the hospital pharmacy’s crisis management team**. It is recommended that this team be composed of **hospital pharmacy management team members reinforced by internal or external staff with specific skills**. | *Minimum staffing numbers; required functions; leaders and their replacements in the event of an absence; role assignment, with or without job descriptions; actions expected from each stakeholder; defining governance of the various stakeholders; defining meeting places; …* | 3 (20) | | 11 (73) | 1 (7) |
| **IV. RECOVERY** | | | | | | |
| **Coming out of a degraded mode of functioning** | | | | | | |
| 38 | For each critical activity and process, the BCP must set **the phases** **for a return to normal activities**. This can be done by means of a **recovery plan** that defines the steps to be taken to recover from a degraded situation, identifying the activities and processes to be resumed according to the situation and available resources. | *A return to normal chemotherapy manufacturing volumes within three days; a return to the normal weekly drug replenishment frequency for every department; etc. Defined recovery times, levels of recovery, and the resources that need to be mobilised for that recovery.* | 2 (13) | | 12 (80) | 1 (7) |
| **Feedback on experiences** | | | | | | |
| 41 | The BCP should provide for the stages in the **analysis of the crisis situation**, including an early operational debriefing (“**hot**” debriefing) and later feedback to the directorate of the hospital with recommendations for improvement. | *Examples of items to analyse: Chronology of events; foreseeable or reducible disruptions; actions already in place to limit their impact; activities affected by the disruption; human, material or financial consequences; actions implemented during the disruption; difficulties encountered; shortcomings identified in the BCP’s ability to handle disruptions; analysis of actions that worked well and those that need improvement; financial impact of the emergency event and its management; planned improvements to the BCP; planned reporting.* | 3 (20) | | 11 (73) | 1 (7) |
| **New statement number** | **Recommendation** | **Examples** | **Median (n = 15)** | **Dissensus** | | **Rewording required** |
| **I. REDUCTION** | | | | | | |
| **Collaboration with partners** | | | | | | |
| 42 | As other **regional hospital pharmacies** are important external partners, the BCP can include specific aspects of **mutual coordination and support** with them. | *Partnership agreements in principle; subcontracting agreements; pooling of human resources (e.g. pharmacists, pharmacy assistants) or material resources (e.g. medicines, manufacturing equipment); ability to take over certain activities in whole or in part (e.g. supply or storage of medicines, production of certain parenteral medicines); etc.* | 8 | | No | No |

N = 15 experts. BCP, business continuity plan; BIA, business impact analysis; IT, information technology. Rephrased elements between Round 1 and Round 2 are highlighted in yellow.

### Experts rating

In Round 1, for each recommendation, experts had to answer the following question by choosing a number on a scale from 1 – 9, as represented bellow : “*How much do you agree with this recommendation?*”

This applied also in Round 2, for each **new** recommendation or recommendation **that has faced dissensus during Round 1**.

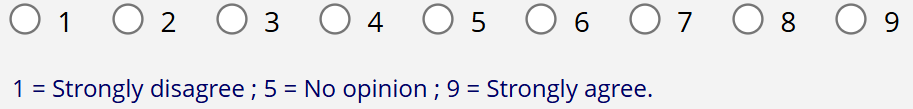

They could also add comments if desired, as represented below :

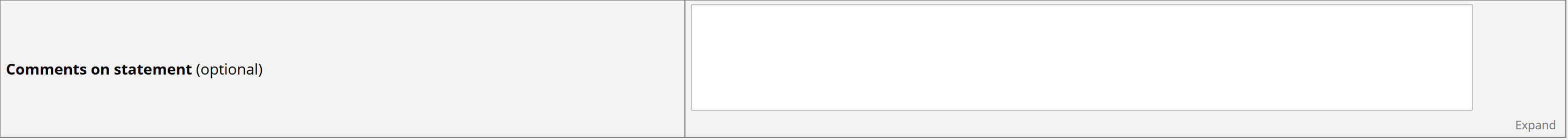

In Round 2, for each **rephrased** recommendation, experts had to indicate their preference between the first and second version of phrasing, as below :

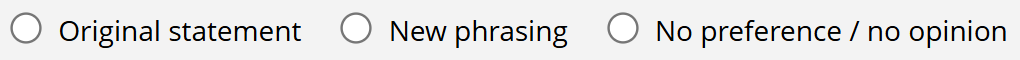
